# Assessment of Workers’ Personal Vulnerability to COVID-19 Using “COVID-AGE”

**DOI:** 10.1101/2020.05.21.20108969

**Authors:** David Coggon, Peter Croft, Paul Cullinan, Anthony Williams

## Abstract

Decisions on fitness for employment that entails a risk of contracting Covid-19 require an assessment of the worker’s personal vulnerability should infection occur. Using recently published UK data, we have developed a risk model that provides estimates of personal vulnerability to Covid-19 according to sex, age, ethnicity, and various comorbidities. Vulnerability from each risk factor is quantified in terms of its equivalence to added years of age. Addition of the impact from each risk factor to an individual’s true age generates their “Covid-age”, a summary measure representing the age of a healthy UK white male with equivalent vulnerability. We discuss important limitations of the model, including current scientific uncertainties and limitations on generalisability beyond the UK setting and its use beyond informing assessments of individual vulnerability in the workplace. As new evidence becomes available, some of these limitations can be addressed. The model does not remove the need for clinical judgement or for other important considerations when managing occupational risks from Covid-19.

## Background

As countries adapt to the longer-term challenges of Covid-19, doctors increasingly will be asked to advise on the fitness for work of patients who might be unusually vulnerable to the disease because of their age, ethnicity and/or comorbidities. The risk of contracting Covid-19 through work will depend on the potential for close proximity to people who could be carrying the infection, or for contact with material contaminated by the virus; the effectiveness of any measures to reduce transmission (such as barriers or personal protective equipment); and local prevalence of the disease at the time. More important for the individual, however, is the risk of serious illness as a consequence of Covid-19, which will depend also on his/her personal vulnerability should infection occur.

Early in the course of the Covid-19 epidemic, the UK government issued guidance on vulnerability from comorbidities^1, 2^. Necessarily, that advice drew on limited empirical evidence from other countries, and extrapolation from experience with other respiratory infections. More recently, others have published guidance on risk reduction for healthcare workers^3^ and the broader management of return to work in the face of health risks from Covid-19^4^. Neither document attempted to quantify risks from specific comorbidities, but research is now emerging that allows more detailed and reliable assessment of vulnerability to Covid-19. In these circumstances, we judged it timely to analyse evidence on risk factors for mortality from the disease, and apply the findings in a risk model that could be used to estimate personal vulnerability. The model is intended principally to assist decisions on occupational placement of workers in the UK. Over time it can be updated and refined as relevant new data become available.

The online resource is now operational, and we here summarise the methods that we used, the structure of the risk model, and our initial findings. Further detail can be found on the project website^5^.

### Methods and sources of evidence

Our aim was to assess and compare risks of fatality in people who contract SARSCov-2 infection, according to their age, sex, ethnicity, smoking habits, and various comorbidities. In preliminary searches of the published literature, no evidence could be found on risks of fatality in representative samples of people infected by the virus (including those with asymptomatic infection). However, analyses of mortality from Covid-19 in the general population could be expected to provide good proxy measures of relative risk, provided the likelihood of contracting infection did not vary importantly according to the risk factors under consideration (as might occur, for example, because of selective shielding by people with certain comorbidities). In addition, estimates of risk might be possible if data could be found on fatality rates by comorbidity in patients admitted to hospital because of Covid-19, and then combined with information about the prevalence of the same comorbidities in hospitalised Covid-19 patients as compared with the general population.

Because of the urgency to improve on earlier advice, we initially sought reports that would provide the strongest evidence relevant to the UK, and did not attempt systematically to search for, and review, all published evidence that might bear on the risks that we were trying to characterise. In this respect, one paper stood out as particularly suited to our purpose.

That report, from the OpenSAFELY (OS) collaborative, presented first results from a cohort study of more than 17 million adults registered with English general practices and followed up from 1 February 2020 to the earlier of death or 25 April 2020^6^. It used multivariate Cox regression to estimate mutually adjusted hazard ratios (HRs) with 95% confidence intervals (CIs) for death in hospital with confirmed Covid-19 (ascertained by linkage to a national notification system) in relation to risk factors determined from pseudonymised individual primary care records. Data on other deaths in the cohort (needed for censoring of follow-up) had been obtained by linkage to records held by the Office for National Statistics (ONS). The report contained a secondary analysis, which censored follow-up at 6 April 2020, allowing exploration of the possibility that HRs for some comorbidities were underestimated in the main analysis because, in response to advice from the UK government at the end of March, people with those diseases had selectively shielded themselves from exposure to infection. As well as sex, age, ethnicity, smoking habits and multiple comorbidities, analyses in the paper adjusted for deprivation (using an index graded to five levels) and for the administrative region of the patient’s general practice (to allow for varying rates of infection in different parts of the country).

This study had unique strengths. It included a substantial proportion of the adult population of England, and was based on more than 5000 deaths attributed to Covid-19. Moreover, information about risk factors came from data recorded before the onset of infection, which reduced the possibility that ascertainment would be biased in relation to the outcome.

Nevertheless, we sought to check the plausibility of its findings, using data from other studies. This was done using four independent sources of information.

1. Published ONS data on mortality from Covid-19 (as the underlying cause of death) by sex and age in England and Wales during March 2020^7^. The death rates make no allowance for effects of comorbidities, the prevalence of which may vary by age, and between men and women. However, they provided a benchmark against which more fully adjusted estimates of relative risk by sex and age could be compared.
2. Published ONS estimates of sex-specific odds ratios for coronavirus-related deaths by ethnic group in England and Wales^8^. These were adjusted for age, geographical region and various other potential confounders, although not for comorbidities.
3. A report from the International Severe Acute Respiratory and Emerging Infection Consortium (ISARIC) study on outcomes, including mortality, in a cohort of 16,749 patients with Covid-19 admitted to hospitals in England, Wales and Scotland during 6 February 2020 to 18 April 2020 (28% of all such admissions nationally during that period) ^9^. Within the cohort, 49% had been discharged, 33% had died, and 17% continued to receive care at the date of reporting. The study reported the prevalence of various pre-existing comorbidities in the cohort, and used multivariate Cox regression to estimate risk of in-hospital death according to age, sex and selected comorbidities.
4. The prevalence of comorbidities by sex and age in samples of people (intended to be nationally representative) from recent rounds of the Health Survey for England^10-13^.

Although the Health Survey for England data predated the ISARIC study, were only from England, and did not apply the same diagnostic criteria and methods of ascertainment as ISARIC, we could use them to calculate an approximate predicted prevalence of comorbidities in the ISARIC cohort. Comparison of the observed and expected prevalence then gave an indication, albeit approximate, of the age- and sex-adjusted relative risks of being hospitalised with Covid-19 according to comorbidities. When combined with HRs for death following admission to hospital in the ISARIC study, this allowed approximate estimation of relative risks of mortality from Covid-19 among people with the comorbidity in the general population.

We abstracted HRs for risk factors of interest from the OS report, and where possible checked their plausibility against data from the other sources mentioned above. The relative risks that we then adopted for our risk model are shown in Table 1, together with our qualitative assessments of the strength of evidence (“robustness”) on which estimates for each risk factor are based. In the main, these estimates were the HRs from the full follow-up period in the OS analysis, since they were statistically the most precise. However, in a few instances, where they were lower than the corresponding HR in the OS study from shorter follow-up, and it seemed probable that this might reflect selective shielding, we made appropriate adjustments. Also, findings from the ISARIC study suggested that the HR for chronic heart disease in the OS study might be too low, and that was therefore adjusted up slightly. Two previously suggested determinants of vulnerability (smoking and hypertension) were omitted from the risk model because after allowance for other factors, they appeared not to carry any material increase in risk. Further details of the calculations, rationale, and qualitative estimation of robustness of the relative risks shown in Table 1 are available as a supplementary online file and on the project website^5^.

**Table 1:** Vulnerability from risk factors expressed as equivalence to added years of age. Please note section on Limitations

| Risk factor | Relative risk | Equivalent added years of age | Robustness of risk estimate |
| --- | --- | --- | --- |
| Sex |  |  |  |
| Male | 1 | 0 |  |
| Female | 0.5 | -8 | Robust |
| Ethnicity |  |  |  |
| White | 1 | 0 |  |
| Asian or Asian British | 1.6 | 5 | Moderately robust |
| Black | 1.7 | 6 | Provisional |
| Mixed | 1.6 | 5 | Provisional |
| Other non-white | 1.3 | 3 | Provisional |
| Body mass index (Kg/m<sup>2</sup>) |  |  |  |
| <30 | 1 | 0 |  |
| 30-34.9 | 1.4 | 4 | Provisional |
| 35-39.9 | 1.6 | 5 | Provisional |
| ≥40 | 2.4 | 10 | Provisional |
| Asthma |  |  |  |
| Mild (no requirement for oral corticosteroids in past year) | 1.1 | 1 | Moderately robust |
| Severe (requiring oral corticosteroids in past year) | 1.4 | 4 | Moderately robust |
| Diabetes |  |  |  |
| Controlled (HbA1c<58 mmol/mol) | 1.5 | 4 | Moderately robust |
| Uncontrolled ( HbA1c≥58 mmol/mol) | 2.4 | 10 | Moderately robust |
| No HbA1c measure in last 15 months | 1.9 | 7 | Moderately robust |
| Chronic heart disease | 1.4 | 4 | Provisional |
| Chronic respiratory disease (excluding asthma) | 1.9 | 7 | Moderately robust |
| Chronic kidney disease* | 1.7 | 6 | Moderately robust |
| Non-haematological cancer |  |  |  |
| Diagnosed <1 year ago | 1.6 | 5 | Provisional |
| Diagnosed 1-4.9 years ago | 1.2 | 2 | Provisional |
| Diagnosed ≥5 years ago | 1 | 0 | Provisional |
| Haematological malignancy |  |  |  |
| Diagnosed <1 year ago | 3.5 | 14 | Provisional |
| Diagnosed 1-4.9 years ago | 3.1 | 13 | Provisional |
| Diagnosed ≥5 years ago | 1.9 | 7 | Provisional |
| Liver disease | 1.6 | 5 | Provisional |
| Chronic neurological disease other than stroke or dementia** | 2.5 | 10 | Provisional |
| Organ transplant | 4.3 | 16 | Provisional |
| Spleen diseases† | 1.4 | 4 | Provisional |
| Rheumatoid/lupus/psoriasis | 1.2 | 2 | Provisional |
| Other immunosuppressive condition‡ | 1.8 | 7 | Provisional |

\*Glomerular filtration rate <60mL/min/1.73m<sup>2</sup>, as estimated from the most recent serum creatinine measurement.
\*\*Includes motor neurone disease, myasthenia gravis, multiple sclerosis, Parkinson's disease, cerebral palsy, quadriplegia, hemiplegia, malignant primary brain tumour and progressive cerebellar disease.
†Includes splenectomy, or spleen dysfunction (e.g. from sickle cell disease).
‡Includes HIV, conditions inducing permanent immunodeficiency (ever diagnosed), aplastic anaemia, and temporary immunodeficiency recorded within the past year.

### Covid-age

A notable feature of Covid-19 is that mortality rates, whether or not they are adjusted for comorbidities, increase exponentially with age. Thus, in the OS report, there were adjusted relative risks of approximately 1.0945 and (1.0945)^10^ = 2.5 for increases in age of one and 10 years respectively^6^. In these circumstances, vulnerability from other risk factors can conveniently be expressed in terms of the added years of age that would give an equivalent increase in risk^14^. Table 1 sets out vulnerabilities associated with demographic variables and comorbidities in our risk model, quantified as age equivalents as well as relative risks. If it is assumed that when risk factors are present in combination, their relative risks multiply (this is the normal default assumption in regression analyses such as those described in the OS report), then combined effects can be estimated by summing the age equivalent for each. Moreover, by adding the summed age equivalents to the person’s true age, it is possible to generate a summary measure of personal vulnerability. We have termed this summary measure a person’s “Covid-age”. It represents the age of a healthy white male with equivalent vulnerability (white males being the largest demographic group in the UK workforce). Here are some examples:

- A healthy white woman, aged 40, has a Covid-age of (40-8) = 32 years.
- A white man, aged 45, BMI 36, with COPD has a Covid-age of (45+5+7) = 57 years
- An Asian woman aged 50 with uncontrolled diabetes has a Covid-age of (50 - 8 + 5 + 10) = 57 years.

Absolute risks can be obtained by translating Covid-ages into estimated case-fatality rates. The process is complicated by current uncertainties about the prevalence of asymptomatic infection (for a given fatality rate in diagnosed symptomatic cases, a higher relative prevalence of asymptomatic cases will imply lower overall case-fatality). However, in a report by Ferguson and colleagues^15^, which drew on findings from a study by Verity and colleagues^16^, the case-fatality rate at 40-49 years of age in men and women combined was estimated to be 1.5 per 1000. Assuming a relative risk of 0.5 in women as compared with men (see Table 1), this would imply a case fatality rate in men of 1.5*2/1.5 = 2 per thousand in men aged 40-49 years. Given that some men in this age band will have comorbidities and other risk factors that increase their vulnerability, this fatality rate might correspond to an average Covid-age in the region of 47 years.

### Limitations

Our analysis and risk model have important limitations.

#### Scientific uncertainties

Currently, all our assessments of risk are derived from a single study, albeit with checks on plausibility using other sources. The outcome in that investigation was death in hospital from Covid-19, and did not extend to deaths elsewhere. Data on some risk factors were incomplete (although the extent of missing information was generally small). Although the study was large, its findings, in particular for rarer comorbidities, are liable to statistical uncertainties (confidence intervals for HRs from the OS report are presented in the documentation that accompanies the vulnerability assessments^5^). Risks associated with some comorbidities may have been attenuated by adjustment for deprivation. We have assumed as a first approximation that relative risks from different factors multiply, but that may not always be true. Also, some risk factors may have been associated with differences in exposure to infection, as well as with differences in vulnerability once infection occurred. The scope for such bias should have been reduced by adjustment of HRs for region, and by our taking into account findings from the earlier OS analysis time-point, when little impact would have been expected from selective shielding of patients with comorbidities. However, we cannot rule out a residual bias, which would have tended towards slight underestimation of the effects of some comorbidities.

#### Heterogeneity of comorbidities

A further limitation is the heterogeneity of some categories of comorbidity for which risk estimates are available. For example, chronic pulmonary disease aggregates various disorders, each with a range of severity. In the future, data should emerge that allow evidence-based risk assessments for more specific sub-categories of disease. Meanwhile, clinical judgement should be applied when considering how risks might vary within a broader, aggregated category of comorbidity, taking as a starting point the estimated risk for the category as a whole.

#### Fatality as an outcome

Our assessment of vulnerability uses fatality as an outcome, and does not take into account loss of life-years. An avoidable death in a young person will normally result in much greater loss of life-years than one occurring at older ages.

#### Generalisability

Although the methods that we have employed may be relevant to development of similar models for other populations, our risk model is designed for application specifically to adults in the UK. It was developed to assist decisions on occupational placement of workers in the UK, and, although some of its findings might be of utility for other purposes, it is not intended to inform decisions in clinical care.

## Conclusion

We expect that some of these limitations can be addressed as further evidence becomes available. Meanwhile, we believe that our assessment of vulnerability offers an improvement on what previously has been available. We caution against simplistic rules for decisions based only on the impacts that it estimates. It does not remove the need for clinical judgement, and there are other important considerations when managing occupational risks from Covid-19 – for example, the practicability of different possible control measures, the personal value judgements of the individual worker, and prevailing advice from government (which may be driven by a need to control demands on healthcare services as well as individual risk). With these caveats, we hope that it will prove a useful contribution to decisions about fitness for work during the Covid-9 pandemic in the UK adult population.

DC contributed to data abstraction, analysis and interpretation, wrote the first draft of the paper, and is its guarantor, PCr contributed to identifying key papers, extraction and interpretation of data, and writing. PCu contributed to the discussions around Covid-age and to the manuscript. AW conceived and organised the project and created the Joint Occupational Health Covid-19 Group to oversee, advise and communicate the results.

## Data Availability

All data used is available in the public domain

## Acknowledgements

We thank the members of the Joint Occupational Health Covid-19 Group who stimulated this work and provided valuable constructive comment. Their names are listed on the project website.

## Conflicts of Interest

All authors declare no conflict of interests.

## Notes

### Competing Interest Statement

The authors have declared no competing interest.

### Funding Statement

No funding supported the work presented

